# A Local Enrichment Computational Framework to Identify Disease-Enriched Physical Activity Patterns

**DOI:** 10.64898/2026.10.22.26366260

**Authors:** Hanyu Qian, Chao Wu, Bo Li, Anthony Rosenzweig, Meng Wang

## Abstract

Wearable accelerometers enable continuous monitoring of physical activity (PA), yet conventional summary measures and population-wide clustering approaches may overlook disease-associated behavioral heterogeneity. Here we introduce LocPA, a computational framework that identifies disease-enriched PA patterns through confounder-adjusted activity profiling, similarity network construction, local disease-enrichment testing and community detection. Applying LocPA to 24-hour accelerometry profiles from the UK Biobank, we identified two recurring PA patterns across six diseases spanning cardiometabolic, neurological, psychiatric, renal and cardiovascular conditions: a low-activity pattern characterized by reduced daytime activity and a sleep-disrupted pattern characterized by reduced daytime and elevated nocturnal activity. In hypertension and diabetes, the sleep-disrupted patten was associated with greater disease burden and less favorable clinical and metabolic profiles, differences not consistently captured by conventional PA summary measures. LocPA also identified disease-specific temporal patterns, particularly in Parkinson’s disease. Analyses in an independent National Health and Nutrition Examination Survey (NHANES) cohort supported the recurrence of hypertension- and diabetes-enriched patterns across populations and accelerometry measurements. LocPA provides a flexible, disease-driven approach for uncovering clinically relevant temporal PA patterns, with potential applications in disease risk characterization and personalized behavioral interventions.

## Introduction

The advent of wearable sensors, including smartwatches and accelerometers, has transformed the measurement of human physical activity (PA), enabling continuous and objective monitoring of movement and rest in free-living environments at unprecedented temporal resolution and population scale^1–3^. Wearable-derived measures of moderate-to-vigorous physical activity, light-intensity activity, sedentary behavior and sleep are increasingly used to investigate relationships between daily activity and a broad range of health outcomes^4–6^. Yet these behaviors do not occur independently. Rather, they are temporally organized and interdependent components of a finite 24-hour day, with their intensity, duration and timing jointly characterizing an individual’s daily activity behavior.

The 24-hour PA profile provides a comprehensive representation of these behavioral dynamics by capturing not only the amount and intensity of activity but also its temporal organization and the interplay between activity and rest throughout the day^6^. Recent studies have leveraged such profiles to identify distinct behavioral patterns, including intensity-based patterns ranging from predominantly inactive to highly active lifestyles, as well as timing-based patterns characterized by activity concentrated at different periods of the day. These patterns have been associated with diverse health outcomes. Profiles characterized by higher overall activity are generally associated with lower risks of all-cause mortality, cardiovascular disease, type 2 diabetes and certain cancers, as well as more favorable cardiometabolic and obesity-related traits^7–14^. Associations with the timing of activity, however, are more heterogeneous and appear to vary across health outcomes^15–18^. A comprehensive review by Farrahi and Farhang summarizes this growing body of work on data-driven profiling of accelerometer-derived physical behaviors and their associations with health outcomes in adults^6^.

Most existing 24-hour PA patterns have been identified using unsupervised learning approaches, such as k-means clustering, hierarchical clustering and latent profile analysis, followed by testing the resulting subgroups for associations with clinical outcomes or biomarkers. Although such data-driven population segmentation has revealed substantial heterogeneity in daily activity patterns, these approaches are designed primarily to capture dominant sources of variation in PA profiles by maximizing within-group similarity and between-group differences, rather than to identify activity patterns specifically associated with disease. Consequently, the resulting patterns may not align with disease states or clinically meaningful differences in disease burden. Moreover, dominant variation in wearable-derived PA may reflect demographic, physiological and behavioral factors—including age, sex and circadian preference—that can obscure subtle activity signatures associated with disease. Disease-associated heterogeneity may therefore remain undetected when it does not coincide with the major axes of variation in the overall population.

A computational framework that directly identifies disease-enriched PA patterns within heterogeneous populations is therefore needed. Instead of first partitioning individuals according to overall variation in their activity profiles and subsequently testing the resulting groups for disease associations, a disease-informed approach could directly leverage disease information to identify local PA patterns enriched among individuals with a particular condition while accounting for major sources of non-disease variation. Importantly, a single disease may be associated with multiple, potentially divergent PA patterns, reflecting heterogeneity in disease presentation, severity or behavioral response. Identifying such localized patterns could uncover disease-associated behavioral signatures that are obscured by global population-level clustering or conventional PA summary measures, enabling finer population stratification and expanding the utility of wearable-derived PA data for disease characterization, monitoring and precision health.

Here, we develop LocPA, a local enrichment framework for identifying robust and comprehensive disease-enriched PA patterns from accelerometer data. LocPA integrates unsupervised neighborhood discovery with localized supervised learning on a similarity graph constructed from confounder-adjusted PA residual profiles. The unsupervised component systematically explores local neighborhoods across the heterogeneous PA landscape, whereas localized supervised learning evaluates whether each neighborhood is selectively enriched for the disease of interest. We applied LocPA to the UK Biobank^19,20^ across a broad range of diseases and used the National Health and Nutrition Examination Survey (NHANES)^21,22^ for external validation and comparison. LocPA identified distinct PA profiles associated with elevated disease prevalence or incidence, revealing that individual diseases can be associated with multiple, divergent PA patterns that may not be captured by population-level clustering or conventional PA summary measures. For hypertension and diabetes, these divergent patterns were associated with differences in disease severity, revealing clinically relevant heterogeneity among individuals with the same condition. We further identified distinct PA signatures associated with Parkinson’s disease, supporting the potential of wearable-derived activity pattens as non-invasive behavioral biomarkers. LocPA demonstrated robust performance across diverse diseases and identified consistent disease-associated PA patterns in an independent population, while also revealing population-specific differences. Together, these findings establish LocPA as a generalizable framework for disease-informed PA phenotyping that captures clinically meaningful behavioral heterogeneity, enables finer population stratification and advances the use of wearable-derived PA profiles for precision health.

## Results

### Overview of the LocPA Workflow

LocPA takes 24-hour PA profiles as input and first accounts for potential confounding effects of age, sex and body mass index (BMI), as well as nonlinear interactions between age and time of day, using generalized additive models (GAMs)^23,24^ (Fig. 1a,b and Methods). The resulting residual profiles, hereafter referred to as PA residual profiles, are used for subsequent analyses. A residual of zero indicates activity equal to the expected level for an individual of the same age, sex and BMI at a given time of day, whereas positive and negative residuals indicate higher and lower activity than expected, respectively. LocPA next constructs a PA similarity network from the residual profiles using k-nearest neighbors (k-NN) ^25^ based on Euclidean distance and evaluates local enrichment for the disease of interest (Fig. 1c and Methods)Click or tap here to enter text.. To reduce the multiple-testing burden while retaining coverage of the study population, LocPA uses a greedy set-cover algorithm^26^ to select a subset of representative participants as seeds, with each seed defining a local neighborhood in the PA similarity network. Within each seed neighborhood, disease enrichment is evaluated using logistic regression for prevalent disease or Cox proportional hazards regression for incident disease, with adjustment for relevant confounders (Fig. 1d and Methods). Statistical significance across seed neighborhoods is determined using a weighted Benjamini–Hochberg (BH) procedure^27^ that accounts for dependencies arising from overlapping neighborhoods. LocPA then organizes significant disease-enriched seed neighborhoods into higher-level PA patterns. Connections between significant seeds are defined according to the overlap between their corresponding neighborhoods, and the resulting network is partitioned using Louvain community detection^28^ (Fig. 1e and Methods). Clustering hyperparameters are selected using data-driven criteria to distinguish discrete PA patterns while limiting redundant partitioning. Finally, the resulting disease-enriched PA patterns are characterized and compared according to their 24-hour activity profiles and annotated using relevant clinical characteristics and biomarkers (Fig. 1f). This workflow enables LocPA to identify multiple, potentially divergent PA patterns associated with the same disease while preserving local heterogeneity in the underlying population.

**Fig. 1.**
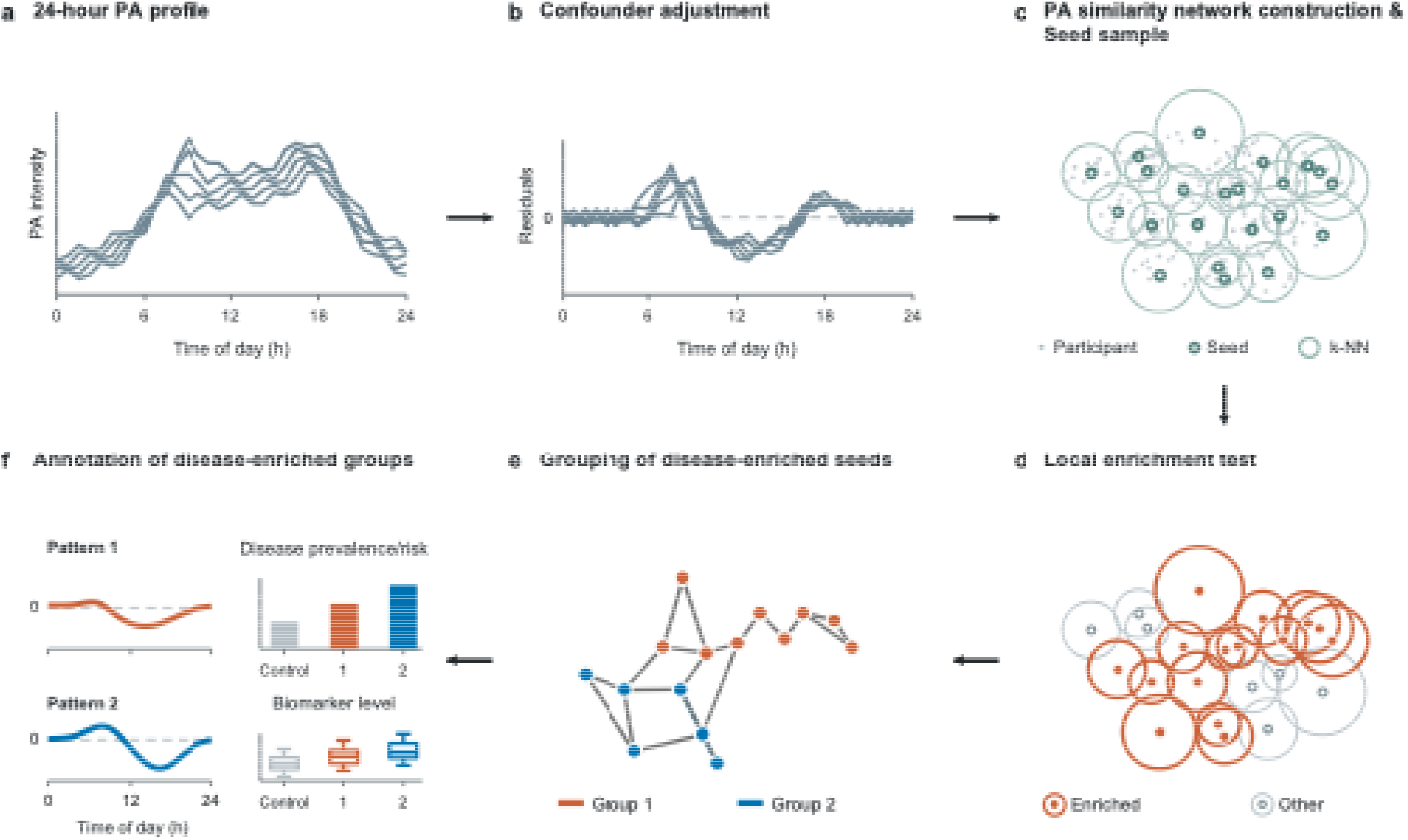
Overview of LocPA. (a) LocPA takes individual 24-hour physical activity (PA) profiles as input. (b) A generalized additive model (GAM) adjusts PA profiles for age, sex and body mass index (BMI) to obtain PA residual profiles. (c) A PA similarity network is constructed from the residual profiles, and representative seed participants are selected to cover the entire cohort while reducing the number of statistical tests. (d) Local enrichment tests assess disease prevalence or future incidence risk within each seed neighborhood, followed by weighted Benjamini–Hochberg (BH) adjustment for multiple testing. (e) Disease-enriched seeds are grouped using Louvain community detection algorithm. (f) The resulting disease-enriched groups are characterized by disease prevalence or incidence risk and associated clinical markers.

### LocPA Reveals Hypertension-Enriched PA Patterns in the UK Biobank

We first applied LocPA to prevalent hypertension, which affects approximately 33% of adults worldwide and 29.6% of UK Biobank participants. The association between PA and hypertension is well established, with higher PA levels consistently associated with a lower risk of hypertension ^29,30^. Consistent with these findings, participants with prevalent hypertension exhibited lower average acceleration than normotensive participants (Supplementary Fig. 1a). However, this population-level difference masked substantial heterogeneity in PA behaviors. Even among participants with hypertension matched for age, sex and BMI, 24-hour PA profiles varied considerably (Supplementary Fig. 1b,c), suggesting that population-level summaries alone may not adequately capture the diverse PA patterns associated with hypertension. LocPA was designed to disentangle this heterogeneity by identifying distinct hypertension-enriched PA patterns rather than relying on average activity levels or predefined population-wide activity profiles.

Following the LocPA workflow (Methods), we generated 24-hour PA residual profiles by adjusting for age, sex and BMI. Using the resulting PA similarity network, we selected 1,212 representative seed participants whose neighborhoods collectively covered all 80,532 participants (Fig. 2a). Of these, 67 seed neighborhoods exhibited significantly elevated hypertension prevalence compared with participants outside the corresponding neighborhoods (weighted BH-adjusted *p* < 0.05 and positive regression coefficients; Fig. 2b, Supplementary Table 2). We further grouped these hypertension-enriched seed PA patterns into two distinct clusters comprising 25 and 42 seeds, respectively (Fig. 2c), using Louvain community detection on the seed-neighborhood overlap network with an overlap threshold of *τ* = 0.01 and a data-driven selected resolution parameter of *γ* = 0.65.

**Fig. 2.**
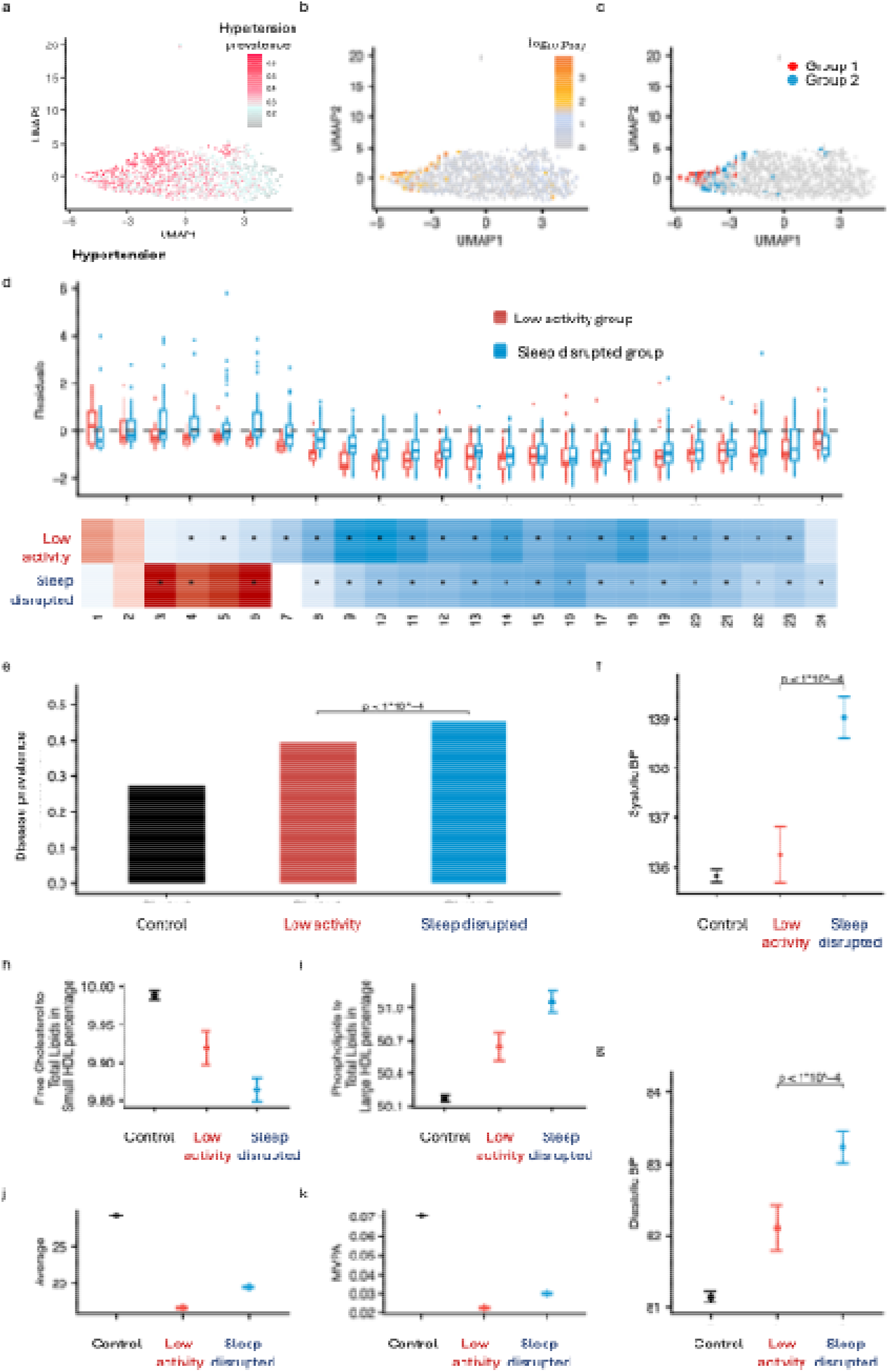
Local enrichment analysis for hypertension prevalence. (a–c) Uniform manifold approximation and projection (UMAP) of seed participants based on PA residual profiles, colored by hypertension prevalence within each seed neighborhood (a), weighted Benjamini– Hochberg (BH)-adjusted p-values from logistic regression (b), and community assignments obtained using Louvain community detection on the seed–seed network (c). (d) Boxplots of hourly PA residuals for seeds in the two identified PA pattens (low-activity and sleep-disrupted; top), and heatmap of mean hourly PA residuals for each pattern (bottom). Asterisks indicate significant deviations from zero (BH adjusted p < 0.05). (e) Hypertension prevalence across the control, low-activity and sleep-disrupted groups, with the highest prevalence observed in the sleep-disrupted group. (f–i) Comparisons of systolic blood pressure (f), diastolic blood pressure (g), Free cholesterol to total lipids in small HDL particles **(h)** and phospholipids to total lipids in large HDL particles **(i)** showing monotonic trends from the control to the low-activity and sleep-disrupted groups. (j,k) Comparisons of average PA intensity (j) and the proportion of time spent in moderate-to-vigorous physical activity (MVPA; k) across the three groups, neither of which follows the ordering of hypertension prevalence. In (f–k), dots represent means, and error bars indicate 95% confidence intervals.

To characterize the two clusters, we examined the 24-hour PA residual profiles of their seed participants and tested for significant deviations from zero, representing the expected PA levels after confounder adjustment (Fig. 2d, Supplementary Table 3). Both clusters exhibited significantly reduced daytime activity relative to expectation (BH adjusted^31^ *p* < 0.05), consistent with the established association between lower PA levels and hypertension. However, their overnight activity patterns differed markedly. The first cluster (25 seeds) showed significantly reduced activity from 04:00 to 23:00, with no significant deviations from expectation during the remaining nighttime hours. We therefore designated this cluster the low-activity group. In contrast, the second cluster (42 seeds) exhibited reduced daytime activity but significantly elevated activity approximately between 03:00 and 06:00, potentially reflecting sleep fragmentation or disruption. We therefore designated this cluster the sleep-disrupted group.

As expected, both hypertension-enriched groups exhibited higher hypertension prevalence than the control group, defined as participants who were not included among the nearest neighbours of any hypertension-enriched seed. Notably, prevalence was significantly higher in the sleep-disrupted group than in the low-activity group (45.4% versus 39.6%; *p* < 1 × 10□□; Fig. 2e). To further characterize their clinical differences beyond hypertension diagnoses, we examined measured blood pressure levels. Both systolic and diastolic blood pressure were significantly higher in the sleep-disrupted group than in the low-activity group (both *p* < 1 × 10□□; Fig. 2f,g, Supplementary Table 4), whereas the control group exhibited the lowest levels. We further compared 248 plasma metabolic biomarkers ^32^ across the control and two hypertension-enriched groups. Three metabolic markers showed significant differences between the low-activity and sleep-disrupted groups and consistent monotonic trends across the control, low-activity, and sleep-disrupted groups (Method, Supplementary Table 6). These included free cholesterol in very large HDL particles, the ratio of free cholesterol to total lipids in small HDL particles, and the ratio of phospholipids to total lipids in large HDL particles. The free cholesterol-to-total-lipids ratio in small HDL particles progressively decreased across the three groups (Fig. 2h), whereas the phospholipid-to-total-lipids ratio in large HDL particles showed the opposite pattern (Fig. 2i). Together, these findings suggest that the sleep-disrupted PA pattern is associated with a more adverse hypertension-related clinical phenotype than the low-activity pattern. Interestingly, conventional PA summary measures associated with hypertension, including average acceleration and the proportion of time spent in moderate-to-vigorous physical activity (MVPA) ^33,34^, did not exhibit corresponding monotonic trends across the three groups (Fig. 2j,k, Supplementary Table 5). These findings highlight the importance of considering the temporal distribution of PA beyond aggregate activity measures and demonstrate the potential of LocPA disease-driven approach to identify distinct PA patterns associated with different levels of hypertension burden.

### LocPA Reveals Diabetes-Enriched PA Patterns in the UK Biobank

We next applied LocPA to prevalent diabetes, a metabolic disorder with well-established associations with PA^35,36^. Diabetes prevalence in the UK Biobank cohort was 3.6%, approximately eightfold lower than that of hypertension. This lower prevalence results in fewer cases within individual seed neighborhoods, potentially reducing statistical power to detect local disease enrichment. Diabetes therefore provides an opportunity to evaluate LocPA ability to identify disease-associated PA patterns under conditions of relatively sparse disease occurrence.

Following the same LocPA workflow, we selected 1,256 representative seed participants whose neighborhoods collectively covered all 85,104 participants (Supplementary Fig. 3a). Of these, 59 seed neighborhoods exhibited significantly elevated diabetes prevalence (weighted BH-adjusted *p* < 0.05 and positive regression coefficients; Supplementary Fig. 3b). Louvain community detection further identified two clusters comprising 23 and 36 seeds, respectively, at the selected resolution of *γ* = 0.75 (Supplementary Figs. 3c and 4).

Remarkably, the two clusters exhibited PA patterns similar to those identified for hypertension. The first cluster (23 seeds) showed significantly reduced daytime activity without substantial deviations from expected activity overnight, corresponding to the low-activity patten. The second cluster (36 seeds) also exhibited reduced daytime activity but showed significantly elevated activity during the early morning hours, corresponding to the sleep-disrupted patten (Fig. 3a, Supplementary Table 7). As observed for hypertension, the sleep-disrupted group exhibited a greater disease burden, with higher diabetes prevalence than the low-activity group (10.6% versus 8.7%, *p* = 0.0065, Fig. 3b). To further characterize their metabolic differences, we examined glycated hemoglobin (HbA1c), a marker of long-term glycaemic control. HbA1c levels were significantly higher in the sleep-disrupted group than in the low-activity group (*p* < 1 × 10□□; Fig. 3c, Supplementary Table 8), while BMI showed a similar increasing trend across the groups (Fig. 3d). Among 248 plasma metabolic biomarkers, 49 showed significant differences between the diabetes-enriched PA pattens and exhibited consistent monotonic trends across the control, low-activity, and sleep-disrupted groups (Supplementary Table 10). These findings further support distinct metabolic profiles associated with the two diabetes-enriched PA pattens.

**Fig. 3.**
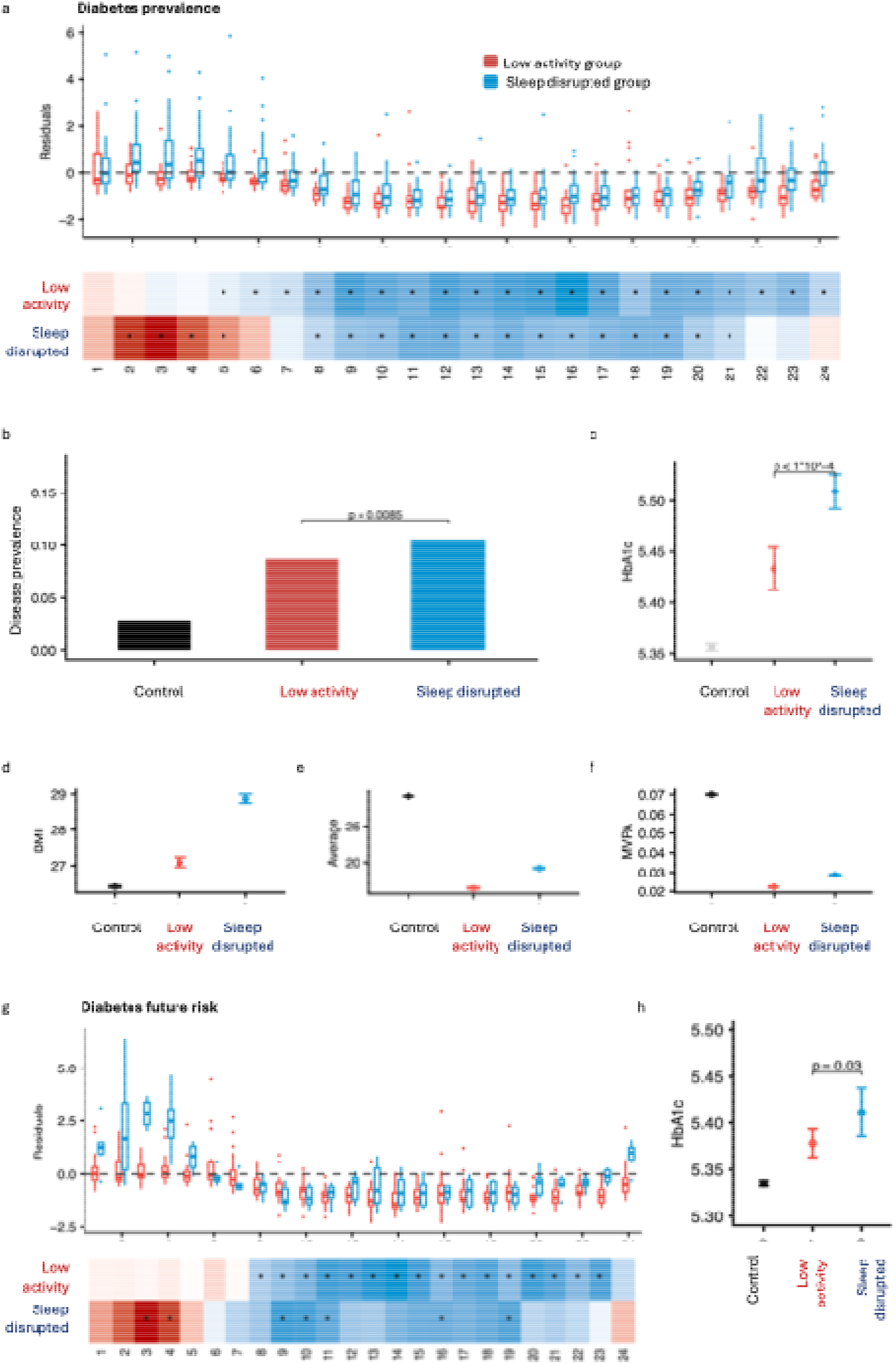
Local enrichment analysis for diabetes prevalence and incidence. (a) Representative 24-h physical activity (PA) profiles for the two identified diabetes-enriched pattens (low-activity and sleep-disrupted; top), and heatmap of mean hourly PA residuals for each pattern (bottom). Asterisks indicate significant deviations from zero (BH adjusted *p* < 0.05). (b) Diabetes prevalence across the control, low-activity and sleep-disrupted groups, with the highest prevalence observed in the sleep-disrupted group. (c,d) Comparisons of glycated hemoglobin (HbA1c; c) and body mass index (BMI; d) across the three groups, with the sleep-disrupted group showing a less favorable metabolic profile. (e,f) Comparisons of average PA intensity (e) and the proportion of time spent in moderate-to-vigorous physical activity (MVPA; f) across the three groups, neither of which follows the ordering of diabetes prevalence. (g) Representative 24-h PA profiles for the two patterns enriched for incident diabetes, recovering the low-activity and sleep-disrupted pattens identified in the prevalence analysis. (h) Comparison of HbA1c across the three groups in the incident diabetes analysis, with higher levels in the sleep-disrupted group than in the low-activity group, consistent with the ordering observed in the prevalence analysis. In (c–f,h), dots represent means, and error bars indicate 95% confidence intervals.

In contrast, conventional PA summary measures did not reflect the ordering of diabetes burden across the groups. Although both diabetes-enriched groups exhibited lower average acceleration and a smaller proportion of time spent in MVPA than controls, both measures were higher in the sleep-disrupted group than in the low-activity group (Fig. 3e,f, Supplementary Table 9), despite its higher diabetes prevalence and HbA1c levels. Together, these findings demonstrate that LocPA identifies distinct diabetes-enriched PA patterns that are also observed in hypertension, with the sleep-disrupted patten consistently associated with a more adverse cardiometabolic profile. The discordance between conventional PA summaries and disease burden further highlights the importance of capturing temporal activity patterns beyond overall activity levels.

We further applied LocPA to incident diabetes to investigate whether distinct PA patterns were associated with future disease risk. After excluding participants diagnosed with diabetes before the accelerometer monitoring index date (Methods), 84,451 participants remained, of whom 2,448 (2.9%) developed diabetes during a median follow-up of 3.88 years. Following the LocPA workflow, we selected 1,250 representative seeds whose neighborhoods collectively covered the cohort. Cox proportional hazards regression^37^ identified 25 seed neighborhoods significantly associated with increased incident diabetes risk (weighted BH-adjusted *p* < 0.05). This number was lower than that identified in the prevalence-based analysis, potentially reflecting the lower event rate and reduced statistical power. Nevertheless, the disease-enriched seeds formed two distinct clusters exhibiting the previously identified low-activity and sleep-disrupted pattens (Fig. 3g, Supplementary Table 11). HbA1c levels also followed the same ordering as incident diabetes risk across the groups (Fig. 3h, Supplementary Table 12). These findings suggest that the two PA pattens are associated not only with prevalent diabetes but also with subsequent disease risk, supporting the relevance of LocPA-derived temporal activity patterns for identifying individuals at elevated risk of developing diabetes.

### Shared and Disease-Specific PA Patterns Across Conditions

We next investigated whether disease-enriched PA patterns were shared across conditions or exhibited disease-specific characteristics. We extended LocPA to four additional diseases spanning neurological, psychiatric, renal and cardiovascular domains: Parkinson’s disease (PD), depression, renal disease and heart failure. Disease prevalence ranged from 0.19% to 8.8% in the UK Biobank (Supplementary Table 2), extending our analysis to conditions substantially rarer than hypertension and diabetes.

Across the six diseases, we identified two recurring disease-enriched PA patterns. The first, termed the low-activity patten, was characterized by persistently reduced daytime activity with minimal deviations from expected activity overnight (Fig. 4a). The second, termed the sleep-disrupted patten, exhibited similarly reduced daytime activity but elevated nocturnal activity (Fig. 4c). To assess the consistency of these patterns across diseases, we performed pairwise permutation tests ^38^ comparing the area between mean PA residual curves (Methods). The low-activity patten was highly consistent across diseases, with significant differences in only two of the 15 pairwise comparisons (PD versus hypertension and PD versus depression; BH adjusted *p* < 0.05, Fig. 4b, Supplementary Table 15). In contrast, the sleep-disrupted patten exhibited greater heterogeneity, with seven significant pairwise differences (Fig. 4d, Supplementary Table 16). Notably, the sleep-disrupted pattern associated with depression differed significantly from those of all five other diseases. Significant differences were also observed between hypertension and diabetes, and between heart failure and renal disease. Although all six diseases shared the characteristic combination of reduced daytime and elevated nocturnal activity, depression exhibited a distinct temporal profile, with the greatest nocturnal activity elevation occurring between 00:00 and 03:00 and comparatively modest daytime activity reductions (Fig. 4e, Supplementary Table 13). These findings suggest that, despite shared broad characteristics, the timing and magnitude of sleep-disrupted PA patterns vary across diseases.

**Fig. 4.**
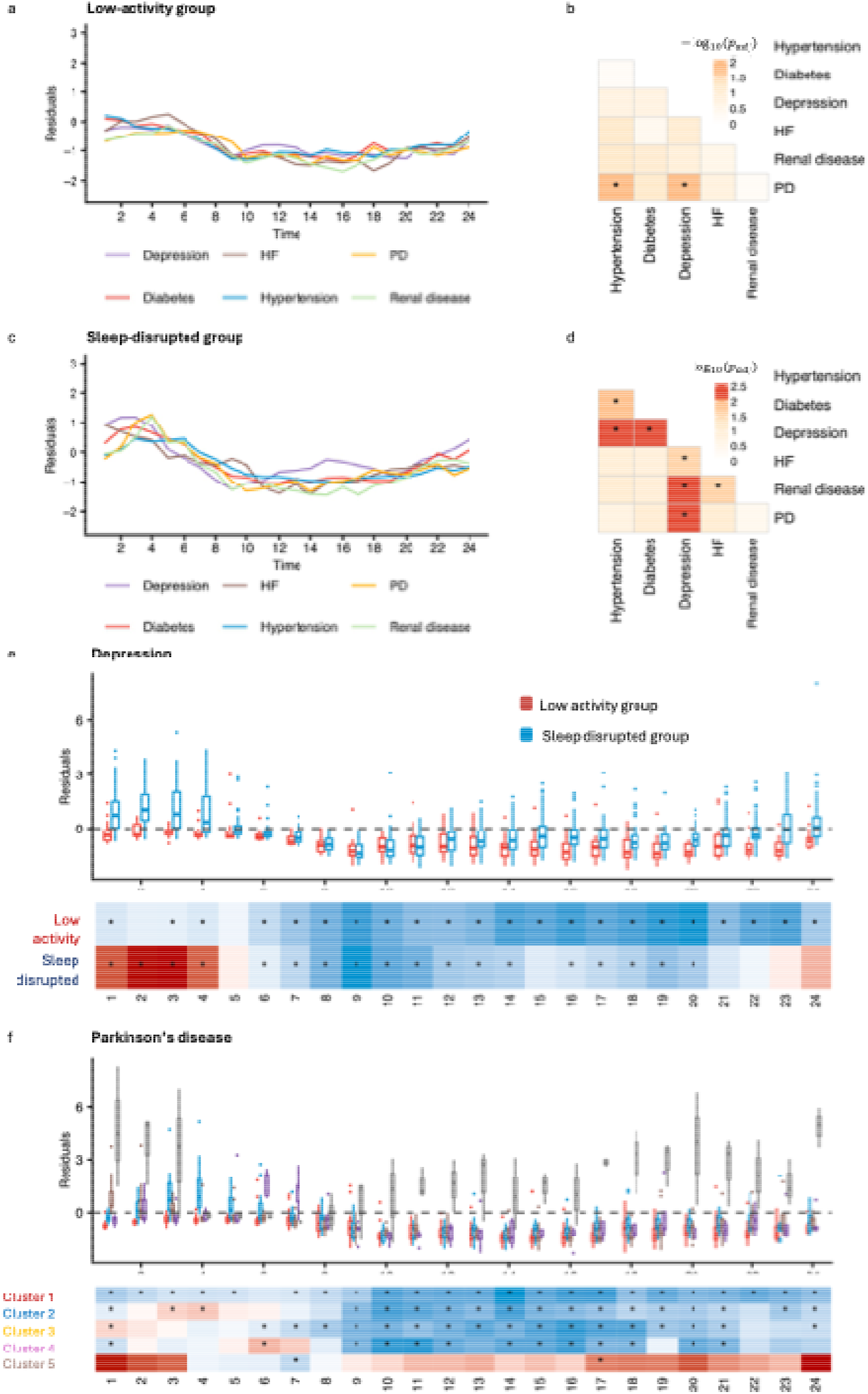
Comparison of disease-enriched PA patterns across diseases. (a,b) Mean hourly PA residual profiles for the low-activity pattern across six diseases (a), showing broadly similar temporal patterns, with significant differences in only 2 of 15 pairwise disease comparisons assessed using permutation tests (b). (c,d) Mean hourly PA residual profiles for the sleep-disrupted pattern across six diseases (c), showing greater heterogeneity, with significant differences in 7 of 15 pairwise comparisons, including all five comparisons involving depression (d). Heatmaps in b and d display −log□□-transformed Benjamini–Hochberg (BH)-adjusted *p*-values from permutation tests; asterisks indicate BH-adjusted p < 0.05. (e) Boxplots of hourly PA residuals for seeds belonging to the low-activity and sleep-disrupted patterns identified in depression (top), and heatmap of mean hourly PA residuals for each patten (bottom). (f) Boxplots of hourly PA residuals for seeds belonging to the five PD-enriched PA patterns (top), and heatmap of mean hourly PA residuals for each pattern (bottom). Cluster 5 exhibits a distinct profile characterized by activity above expected levels throughout much of the day, with particularly pronounced elevations during the evening and nighttime hours. PD, Parkinson’s disease; HF, heart failure.

Beyond these recurring pattens, LocPA also identified disease-specific PA patterns. For PD, five distinct PA patterns were associated with elevated disease prevalence (Fig. 4f, Supplementary Table 14). Notably, the fifth cluster exhibited activity above cohort-expected levels throughout much of the day, with particularly pronounced elevations during the evening and nighttime hours, rather than a uniform increase in overall activity. This pattern remained distinct across a broad range of Louvain resolution parameters (*γ* from 0.5 to 1.2; Supplementary Fig. 5). As the resolution decreased, the other clusters progressively merged, whereas this cluster remained separate, suggesting that it represents a distinct structure within the seed-overlap network rather than an artifact of the selected resolution. The pronounced evening and nocturnal activity elevations may reflect altered daily activity rhythms rather than greater overall PA. However, the small number of contributing seeds warrants cautious interpretation.

### Cross-Cohort Comparison of LocPA-Identified Disease-Enriched PA Patterns

To evaluate the generalizability of LocPA, we applied the framework to hypertension and diabetes in an independent cohort from NHANES ^21,22^. These two diseases were selected because they were assessed in both NHANES and UK Biobank, enabling cross-cohort comparisons. The NHANES accelerometry substudy included 7,707 participants (Methods), substantially fewer than the UK Biobank cohort. To account for differences in population composition, we additionally adjusted for ethnicity, alongside age, sex and BMI. Given the smaller sample size and potentially reduced statistical power, we adopted an exploratory significance threshold for weighted BH-adjusted *p* < 0.35 to identify candidate disease-enriched neighborhoods.

For hypertension, 12 seed neighborhoods met the enrichment criterion (adjusted *p* < 0.35 and nominal *p* < 0.015), and Louvain community detection identified two PA patterns at the selected resolution of *γ* = 0.9 (Fig. 5a, Supplementary Table 17). The first exhibited reduced daytime activity with minimal deviations from expected activity overnight, corresponding to the low-activity pattern. The second was characterized by elevated nocturnal activity, potentially reflecting disrupted sleep or rest–activity rhythms, corresponding to the sleep-disrupted pattern. Both patterns broadly resembled those identified in UK Biobank, with cross-cohort correlations of 0.771 for the low-activity pattern and 0.598 for the sleep-disrupted pattern.

**Fig. 5.**
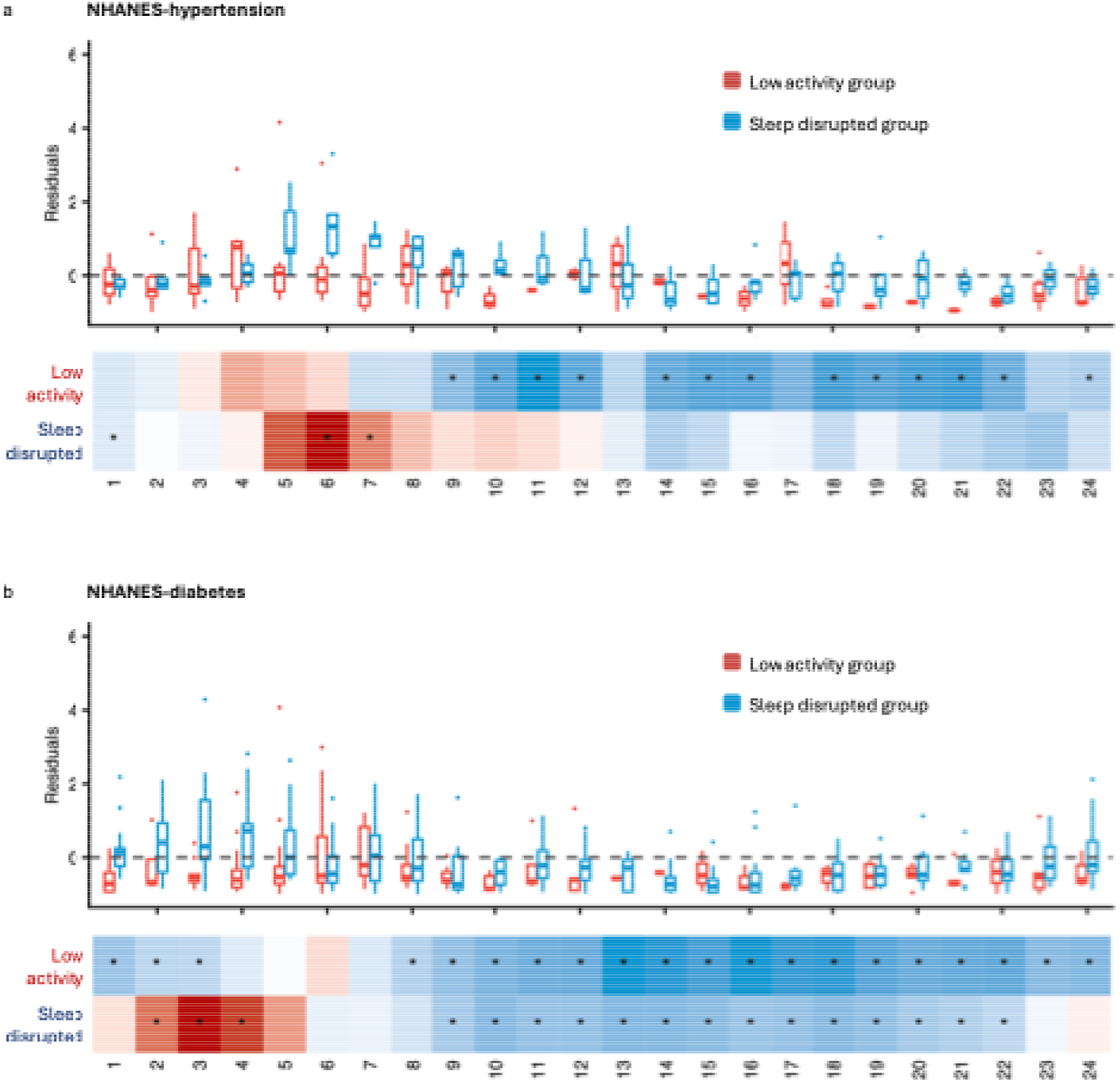
Local enrichment analysis of hypertension and diabetes in the NHANES cohort. (a,b) Boxplots of hourly PA residuals for seeds in the low-activity and sleep-disrupted patterns identified for hypertension (a; top) and diabetes (b; top), with corresponding heatmaps of mean hourly PA residuals for each disease (bottom). Asterisks indicate significant deviations from zero. Cross-cohort correlations between mean hourly PA residual profiles in NHANES and UK Biobank were r = 0.771 for the low-activity pattern and r = 0.598 for the sleep-disrupted pattern in hypertension. For diabetes, the corresponding correlations were r = 0.753 and r = 0.965, respectively.

Similar findings emerged for diabetes, for which 34 seed neighborhoods met the enrichment criterion. Louvain community detection identified two PA patterns at the selected resolution of *γ* = 0.85 (Fig. 5b and Supplementary Table 18). The low-activity pattern exhibited reduced activity primarily during daytime hours, whereas the sleep-disrupted pattern was characterized by elevated nocturnal activity. Both diabetes-enriched PA pattern showed similar temporal profiles across NHANES and UK Biobank, with cross-cohort correlations of 0.753 for the low-activity pattern and 0.965 for the sleep-disrupted pattern. The particularly high correlation for the sleep-disrupted pattern indicates strong consistency of this temporal pattern across cohorts. These findings provide additional support for the applicability of LocPA across populations with different demographic compositions and accelerometry measurements.

## Discussion

In this study, we developed LocPA, a computational framework for identifying disease-enriched PA patterns from 24-hour accelerometry profiles. Unlike conventional approaches that summarize PA using aggregate measures or identify population-wide activity patterns without considering disease associations, LocPA integrates confounder adjustment, PA similarity network construction, local disease-enrichment testing and community detection to uncover distinct temporal PA patterns associated with disease. Applying LocPA to the UK Biobank, we identified two recurring disease-enriched patterns across six conditions spanning cardiometabolic, neurological, psychiatric, renal and cardiovascular domains: a low-activity patten characterized by reduced daytime activity and a sleep-disrupted patten characterized by reduced daytime and elevated nocturnal activity. Notably, the sleep-disrupted pattern was associated with greater hypertension and diabetes burden and less favorable clinical and metabolic profiles, differences that were not consistently captured by conventional PA summary measures. Beyond these shared patterns, LocPA revealed disease-specific temporal signatures, particularly in Parkinson’s disease, highlighting heterogeneity in disease-associated PA behaviors. The recurrence of similar hypertension- and diabetes-enriched patterns in an independent NHANES cohort further supported the generalizability of the framework across populations and accelerometry measurements.

A notable finding of this study is the identification of two recurring disease-enriched PA patterns across six conditions: low activity and sleep disruption. The low-activity pattern, characterized by reduced daytime activity, is consistent with extensive evidence linking physical inactivity to cardiometabolic disorders and adverse health outcomes. Previous accelerometry studies have also demonstrated that lower overall activity and distinct 24-hour PA trajectories are associated with increased mortality and cardiovascular risk^7,39^. More importantly, we identified a sleep-disrupted pattern characterized by reduced daytime activity accompanied by elevated nocturnal movement, suggesting that the temporal distribution of activity may provide disease-relevant information beyond overall activity levels. This observation is consistent with growing evidence implicating disrupted sleep and rest–activity rhythms in multiple diseases. For example, actigraphy studies have documented reduced daytime activity, increased nocturnal awakenings and altered circadian rhythms in individuals with depression^40^, while sleep disturbances and fragmented rest–activity rhythms are frequently observed in Parkinson’s disease (PD)^41,42^. Interestingly, although the two patterns recurred across diseases, their temporal profiles were not identical, particularly for the sleep-disrupted pattern, suggesting potentially distinct disease-related behavioral or physiological processes. Beyond these shared patterns, LocPA identified a distinctive PD-enriched pattern characterized by elevated activity throughout much of the day, with pronounced increases during evening and nighttime hours. This finding is consistent with previous reports of increased nocturnal motor activity, fragmented rest–activity rhythms and altered circadian regulation in PD^41,42^. Rather than indicating greater beneficial PA, this pattern may reflect abnormal motor activity or disrupted rest–activity regulation. These findings suggest that disease-associated PA alterations encompass both shared behavioral pattens and condition-specific temporal signatures, highlighting the importance of examining when activity occurs, rather than solely how much activity is accumulated.

Beyond identifying distinct disease-enriched PA patterns, LocPA revealed their associations with clinical and metabolic characteristics, highlighting the potential clinical relevance of temporally resolved activity profiles. In both hypertension and diabetes, the sleep-disrupted patten was associated with a greater disease burden than the low-activity patten, accompanied by higher systolic and diastolic blood pressure in hypertension and elevated glycated hemoglobin (HbA1c) and BMI in diabetes. These findings are consistent with previous evidence linking disrupted sleep and rest–activity rhythms to cardiometabolic dysfunction. Experimental studies have shown that sleep restriction and circadian misalignment can impair insulin sensitivity^43^, while epidemiological studies have associated altered rest–activity rhythms, including reduced rhythm amplitude and elevated activity during the usual resting period, with increased risk of type 2 diabetes and cardiovascular outcomes^44–47^. Importantly, the distinct PA patterns identified by LocPA were also accompanied by differences in plasma metabolic biomarkers. In hypertension, these differences were limited to three HDL-related measures reflecting lipid content and composition. In diabetes, a broader metabolic signature was observed, spanning HDL and VLDL particle characteristics, lipid composition, triglyceride-related measures, and fatty acid profiles. Several of the metabolic markers distinguishing the diabetes-enriched PA pattens have previously been associated with diabetes. For example, average VLDL particle diameter progressively increased from the control to the low-activity and sleep-disrupted groups, with the highest level observed in the sleep-disrupted group. Larger VLDL particle size has been associated with higher diabetes risk^48^. In contrast, the proportion of polyunsaturated fatty acids relative to total fatty acids progressively decreased across the three groups and was lowest in the sleep-disrupted group, consistent with previous evidence linking higher PUFA levels to lower diabetes risk^49^. These patterns further support a less favorable metabolic profile in the sleep-disrupted patten. Notably, conventional PA summary measures, including average acceleration and time spent in moderate-to-vigorous PA, did not consistently reflect the observed ordering of disease burden and metabolic characteristics. This discordance suggests that the timing and distribution of activity may provide complementary information to total activity volume for characterizing cardiometabolic health. These findings highlight the potential of LocPA to identify clinically and metabolically distinct subpopulations that may be overlooked by conventional PA assessments.

LocPA offers several conceptual and methodological advantages over conventional unsupervised clustering approaches for identifying disease-associated PA patterns. Traditional population segmentation methods typically cluster individuals based solely on similarities in their PA profiles and subsequently examine disease prevalence or risk across the resulting groups. Because clustering is driven by the overall structure of the PA data rather than disease associations, the resulting clusters may primarily reflect dominant sources of behavioral variation, potentially obscuring subtle but clinically relevant patterns that occur in relatively small subpopulations. An alternative strategy is to cluster PA profiles exclusively among individuals with a given disease. However, this approach identifies heterogeneity within the affected population without directly establishing which patterns are specifically enriched for the disease relative to the broader population. Moreover, it excludes unaffected individuals who may exhibit similar PA patterns, limiting its ability to characterize disease-associated behavioral pattens across the full disease continuum or investigate their associations with future disease risk. In contrast, LocPA integrates disease outcomes directly into pattern discovery through local enrichment testing within a similarity network constructed from the entire population. This disease-driven approach enables the identification of localized PA patterns associated with elevated disease prevalence or incidence without requiring them to form globally distinct clusters. By subsequently grouping enriched neighborhoods with similar activity profiles, LocPA can reveal both shared and disease-specific patterns while preserving heterogeneity that may be overlooked by conventional clustering. Importantly, the framework does not require predefined PA categories or restrict pattern discovery to individuals with diagnosed disease, providing a flexible approach for characterizing disease-relevant behavioral variation across populations.

An additional strength of LocPA is its flexibility and potential generalizability across PA representations, study populations and disease outcomes. Although the present study focused on 24-hour PA profiles summarized at hourly intervals, the framework is not restricted to this temporal resolution or representation. It can, in principle, be extended to finer-resolution activity profiles that capture short-duration behavioral fluctuations, longer-term activity trajectories that characterize day-to-day or weekly variability, or low-dimensional representations learned from raw accelerometry. By constructing similarity networks in these alternative representation spaces, LocPA could identify disease-enriched behavioral patterns that are not readily captured by hourly activity summaries. The framework is also adaptable to different disease outcomes, including prevalent and incident conditions, and potentially continuous clinical phenotypes through appropriate modifications of the local enrichment models. Importantly, our analyses of UK Biobank and NHANES demonstrated the recurrence of low-activity and sleep-disrupted pattens associated with hypertension and diabetes across independent cohorts with different demographic compositions, accelerometry measurements and sample sizes. Nevertheless, the exact temporal profiles differed between cohorts, suggesting that disease-associated PA patterns may exhibit population- and measurement-specific heterogeneity. Such differences could arise from variations in demographic characteristics, behavioral habits, disease characteristics, accelerometer devices, activity quantification methods and data collection protocols. Thus, while the broad disease-enriched pattens may be reproducible across populations, their detailed temporal signatures should not be assumed to be identical. These findings highlight both the adaptability of LocPA and the importance of accounting for cohort-specific characteristics when interpreting and translating disease-associated PA patterns.

Several limitations of this study warrant consideration. First, the statistical power of LocPA depends on cohort size, disease prevalence or incidence, and the number of cases within local neighborhoods. For rare diseases or outcomes with low event rates, limited case counts may reduce the ability to detect significant disease enrichment, potentially leading to missed disease-associated PA patterns. This limitation may be particularly relevant in smaller cohorts or analyses of incident diseases with relatively short follow-up. Second, LocPA relies on several hyperparameters, including the number of nearest neighbors *k u*sed to construct the PA similarity network, the neighborhood-overlap threshold *τ* used to define the seed–seed network, and the Louvain resolution parameter *γ* used for group detection. These parameters influence neighborhood composition, the number of enriched seeds and the granularity of the resulting PA patterns. Although we implemented systematic parameter selection procedures and evaluated the stability of identified patterns, optimal settings may vary across cohorts, disease outcomes and PA representations, necessitating further calibration and sensitivity analyses in future applications. Third, the observational nature of the study precludes causal interpretation of the identified associations. Disease-related symptoms, functional limitations, medications and other unmeasured factors may influence PA behaviors, particularly in analyses of prevalent diseases, making it difficult to distinguish behavioral risk factors from consequences of disease. Although the analysis adjusted for major demographic and anthropometric confounders, residual confounding may remain. Finally, although the recurrence of similar pattens in UK Biobank and NHANES provides initial evidence of cross-cohort generalizability, differences in population characteristics, accelerometry measurements and disease ascertainment may affect the identified patterns. Further validation in larger, more diverse longitudinal cohorts, ideally with complementary sleep, clinical and behavioral measurements, will be important to establish the robustness, interpretability and clinical utility of LocPA.

In conclusion, we developed LocPA, a disease-driven computational framework for identifying disease-enriched physical activity patterns from accelerometry data. Across multiple diseases, LocPA revealed shared and disease-specific temporal patterns associated with distinct clinical and metabolic profiles, with recurring patterns observed across independent cohorts. By capturing disease-relevant behavioral heterogeneity beyond conventional activity summaries, LocPA provides a flexible framework for investigating PA–disease relationships and advancing personalized risk assessment and behavioral interventions.

## Methods

### Study participants

We used the UK Biobank (UKB) as the primary cohort for LocPA development and evaluation. UKB is a large population-based prospective cohort comprising approximately 500,000 participants aged 40–69 years at recruitment ^20^. Between 2013 and 2015, approximately 100,000 participants participated in the UKB accelerometry substudy, wearing wrist-worn triaxial accelerometers that continuously recorded physical activity for up to seven days^5,19^. Accelerometer data were processed according to the established UKB preprocessing and quality-control procedures^5^. Participants were excluded if their accelerometer recordings were of poor quality, had insufficient wear time or could not be adequately calibrated. After quality control, 87,716 participants with valid accelerometer data were included in the analysis. Our study was conducted under UK Biobank application 197947, with data accessed by the University of Michigan. All UKB participants provided written informed consent, and the UK Biobank study received ethical approval from the relevant institutional review boards.

We used the National Health and Nutrition Examination Survey (NHANES) cohort for external validation and cross-population comparison. NHANES is a nationally representative, cross-sectional survey of the US population that collects demographic, health and behavioral data, including accelerometer-based measurements of physical activity over seven consecutive days. We excluded participants younger than 20 years, pregnant participants and those with fewer than four valid days of accelerometer recordings. The final analytic sample comprised 7,701 participants aged 20–79 years (48.7% male) from the 2011–2014 NHANES cycles^21,22^.

### 24-hour PA profile from accelerometer data

Accelerometer-derived physical activity intensity was quantified using the Euclidean norm minus one (ENMO) metric in UKB ^5,50^ and Monitor-Independent Movement Summary (MIMS) units in NHANES ^51^. Although these metrics differ in their computational approaches and measurement scales, both provide measures of movement intensity derived from raw triaxial acceleration signals. For each participant, we constructed a 24-hour PA profile by averaging the corresponding valid intensity measurements within each clock hour across the seven-day monitoring period. PA profiles were analyzed separately within each cohort.

### Disease ascertainment and classification

Disease diagnosis information in UKB was obtained from the first occurrence dataset (Category 1712) ^20^. These records integrate information from primary care, hospital inpatient records, death registry data, and self-reported medical conditions to identify the earliest recorded occurrence of each disease. Diseases were defined using International Classification of Diseases, Tenth Revision (ICD-10) codes, with the corresponding codes for all diseases included in this study provided in Supplementary Table 2.

In UKB, the start of the accelerometer monitoring period was defined as the index date for disease classification and follow-up. Participants with a recorded diagnosis on or before the index date were classified as prevalent cases, whereas those with a first recorded diagnosis after the index date were classified as incident cases. For prevalent disease analyses, participants who developed the corresponding disease only after the index date were excluded. For incident disease analyses, participants with prevalent disease at the index date were excluded, and followup time was calculated from the index date to the first recorded diagnosis or the administrative censoring date of 1 September 2023, whichever occurred first.

In NHANES, disease status was ascertained from self-reported medical history questionnaires. Participants were classified as having a disease if they reported having been told by a healthcare professional that they had the corresponding condition. Because the timing of disease onset relative to accelerometer monitoring could not be consistently established across conditions, NHANES disease outcomes were analyzed exclusively as prevalent disease status and used for external validation of the disease prevalence analyses conducted in UKB.

### Phenotypic characteristics and biomarkers

For UKB participants, demographic, anthropometric and biomarker data were obtained from the UK Biobank assessment records. Age was defined as the time of accelerometer monitoring, and sex was obtained from the UKB participant records. Body mass index (BMI), blood pressure and glycated hemoglobin (HbA1c) were measured during the baseline assessment. In addition, 248 plasma metabolic biomarkers^32^ measured at baseline were included to provide a comprehensive characterization of participants’ metabolic profiles. As the accelerometry substudy was conducted several years after the initial UKB assessment, these baseline phenotypic and biomarker measurements generally preceded accelerometer monitoring.

For NHANES participants, age, sex and BMI were obtained from the corresponding survey cycles in which accelerometer data were collected, reflecting participant characteristics at or near the time of PA monitoring.

### Confounder adjustment

To reduce variation in 24-h PA profiles attributable to demographic and anthropometric characteristics, we used confounder-adjusted PA residuals rather than raw activity intensity for subsequent analyses. Generalized additive models (GAMs) ^23,24^ were used to model the expected hourly PA profile as a function of age, sex, BMI, and nonlinear interactions between age and time of day.

For each target disease, a separate GAM was fitted using participants without the disease of interest to establish the expected PA reference profile. Let *Y*_*it*_ denote the mean activity intensity for participant *i* during hour *h*, where *h =* 1,2, …,24. The model was specified as

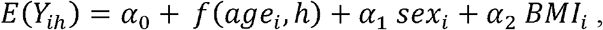

where *f* (*age*_*i*_,*h*) represents a tensor-product smooth of age and time of day, allowing the shape of the 24-h activity profile to vary nonlinearly with age. The tensor-product smooth was constructed using cubic regression spline bases for both time and age, with marginal basis dimensions of 10 and 5, respectively. Sex and BMI were included as parametric confounders.

The fitted model was then used to predict the expected hourly activity intensity for every participant, including both participants with and without the target disease. The confounder-adjusted PA residual was calculated as

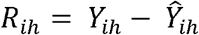

where *Ŷ*_*ih*_ is the activity intensity predicted by the GAM. Each participant was therefore represented by a 24-dimensional residual PA profile, which was used as input for the subsequent analyses.

### PA similarity network construction and seed selection

We constructed a PA similarity network using the *k*-nearest-neighbor (k-NN) algorithm^15^ based on Euclidean distances between confounder-adjusted PA residual profiles. The neighborhood size was set to 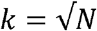, where *N d*enotes the total number of participants, as a heuristic to balance local pattern resolution with sufficient neighborhood size for subsequent statistical enrichment analyses. For each participant *i*, we defined a local neighborhood ℕ_*i*_ consisting of the participant and their (*k* − 1) nearest neighbors in the residual PA profile space.

To reduce the multiple-testing burden while maintaining coverage of the study population, we selected a subset of representative participants, referred to as seeds, using a greedy set-cover algorithm ^26^. At each iteration, the participant whose neighborhood contained the largest number of individuals not yet covered by previously selected seed neighborhoods was selected as the next seed. This procedure was repeated until all participants were covered by at least one selected neighborhood. The resulting set of seed participants, denoted by S, and their corresponding neighborhoods, ℕ_*S∈*S_, were subsequently used for local disease enrichment analyses.

### Local disease-enrichment testing and false discovery rate control

For each disease, we evaluated whether participants within each seed neighborhood ℕ_*S*_ exhibited a higher disease burden than those outside the neighborhood. Prevalent and incident disease outcomes were analyzed separately.

For prevalent disease, logistic regression was used to assess the association between seed-neighborhood membership and disease status. Let *I*_*is*_ indicate whether participant *i* belonged to the neighborhood of seed *s*. The model was specified as

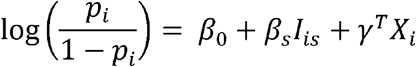

where *p*_*i*_ is the probability of prevalent disease for participant *i, X*_*i*_ includes age, sex and BMI, and *β*_*S*_ quantifies the association between neighborhood membership and disease prevalence after confounder adjustment. Statistical significance was evaluated using two-sided tests, and neighborhoods with a positive coefficient *β*_*S*_ >0 and statistically significant association after multiple-testing correction were considered enriched for prevalent disease.

For incident disease, Cox proportional hazards regression ^37,52^ was used to evaluate the association between seed-neighborhood membership and subsequent disease risk. The hazard function was specified as

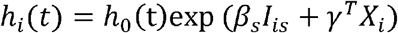

where *h*_*i*_(*t*) is the hazard of disease onset for participant *i* at follow-up time *t* and *h*_0_(*t*) is the baseline hazard function. The coefficient *β*_*S*_ represents the log hazard ratio associated with membership in seed neighborhood ℕ_*S*_. Separate neighborhood-level tests were conducted for each disease.

To account for multiple testing across seed neighborhoods in the PA similarity network, we applied a weighted Benjamini–Hochberg (BH) procedure ^27,53^. The procedure incorporates the local density of tested seeds in the PA residual space, assigning greater weights to seeds in densely populated regions and lower weights to those in sparsely populated regions. Specifically, for each seed *s*, local density was approximated by the reciprocal of the Euclidean distance to its [k/2]th nearest neighbor among the |S| seeds. These density-based weights were used to adjust the enrichment-test *p*-values. The weighted BH procedure was applied at a target false discovery rate of 0.05, and seed neighborhoods with significant adjusted *p*-values and positive regression coefficients were identified as disease-enriched.

### Clustering disease-enriched PA patterns

For each disease, we represented each disease-enriched seed neighborhood by the PA residual profile of its seed participant and grouped seeds according to the overlap between their corresponding neighborhoods. Specifically, we constructed a seed-level adjacency matrix (A), with connections defined by

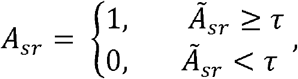

where 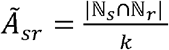 measures the overlap between the neighborhood of seeds *s* and *r*, and *τ* is a predefined threshold.

We applied the Louvain community detection algorithm ^28,54,55^ to the resulting seed network to identify groups of disease-enriched PA patterns. The Louvain resolution parameter *γ* was selected using the average silhouette width, which evaluates the separation and cohesion of the resulting groups. We evaluated *γ* from 0.1 to 2.0 in increments of 0.05, performing 20 independent runs with different fixed random seeds at each resolution to account for stochastic variation in community detection.

Candidate resolutions were considered admissible if at least 80% of seeds were assigned to groups containing three or more seeds, ensuring sufficient coverage by nontrivial groups, and if the number of such groups ranged from two to six, balancing pattern diversity and interpretability. Among admissible resolutions, we retained those whose mean silhouette width was within one standard error of the maximum mean silhouette width across runs, allowing selection of resolutions with near-optimal clustering quality. We then selected the resolution yielding the largest median number of groups to capture greater PA pattern heterogeneity without substantially compromising cluster separation. Ties were resolved by selecting the larger resolution. At the selected resolution, the clustering result from the run with the highest average silhouette width was retained. The same procedure was applied independently to each disease.

### Sensitivity Analyses of LocPA Parameter Settings

We assessed the robustness of LocPA to key hyperparameter choices using hypertension as a benchmark. First, we evaluated the sensitivity of the weighted BH procedure to the choice of neighborhood density weights. We compared weights derived from the distances to the [*k*/2]-th and *k*-th nearest neighbors and found that they were nearly perfectly rank-correlated (Spearman’s *ρ* = 0.99), resulting in identical sets of significantly enriched seeds. We therefore used the distance to the [*k*/2]-th nearest neighbor to calculate weights in the primary analysis. Second, we examined the sensitivity of community detection to the neighborhood-overlap threshold *τ* used to construct the seed–seed network. Varying *τ* from 0.0075 to 0.015 yielded cluster assignments that were largely consistent with those obtained using the selected threshold *τ* = 0.01, with adjusted Rand indices ranging from 0.67 to 1.00. Finally, we compared average silhouette width and the Calinski–Harabasz index as criteria for selecting the resolution parameter *γ* in Louvain community detection. Both criteria yielded the same number of clusters and similar disease-enriched patterns (Supplementary Fig. 6).

### PA pattern comparison

To assess differences in PA patterns between clusters, we performed a permutation test ^38^ based on the area between their mean residual PA profiles. For clusters a and b, containing *n*_*a*_ and *n*_*b*_ participants, respectively, the mean residual PA profiles were denoted by 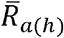 and 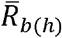, where *h* represents the hour of day. The observed area between curves (ABC) was calculated as

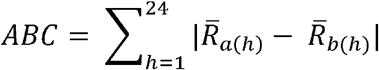

Larger ABC values indicate greater overall differences between the two PA patterns. To assess statistical significance, we generated an empirical null distribution by pooling seed participants from both clusters and randomly permuting their cluster labels while preserving the original cluster sizes *n*_*a*_ and *n*_*b*_. For each permutation, we recalculated the mean residual PA profiles and the corresponding ABC statistic. This procedure was repeated B = 1,000 times. The permutation *p*-value was calculated as the proportion of permuted ABC statistics greater than or equal to the observed ABC, i.e.,

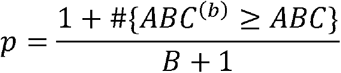

### Plasma metabolomic comparison

To identify plasma metabolic biomarkers that differed across PA groups, we performed a two-stage analysis. First, we compared metabolite levels between the combined disease-enriched groups and the control group, applying Bonferroni correction^56^ for multiple testing (*p* < 0.05/M, where *M* denotes the total number of measured metabolites). Second, metabolites significant in the first stage were further compared between the disease-enriched PA groups using a Bonferroni-corrected threshold of *p* < 0.05/*M*_*s*_, where *M*_*s*_ denotes the number of metabolites identified in the first stage. Finally, we examined whether metabolites significant in both stages exhibited monotonic trends across the control, low-activity and sleep-disrupted groups.

### Computing hardware and software

All statistical analyses were performed using R (version 4.5.0). GAMs were fitted using the **mgcv** package^23^, nearest neighbors were identified using the **RANN** package^25^, and LCD was implemented via the **igraph** package^54,55^. All source code for the local enrichment analysis framework is publicly available on GitHub.

## Data Availability

All data produced in the present study are available upon reasonable request to the authors

## Data and code availability

All UK Biobank data are accessible via application to the UK Biobank. All analyses and visualization were implemented in R (v4.5.0). Scripts are available on GitHub: https://github.com/mengwang-lab/LocPA.

## Acknowledgements

M.W., and H.Q., was supported by M.W.’s startup fund from the Institute for Heart and Brain Health. This work was supported by grants the NIH R35 HL155318 to A.R. and the American Heart Association (AHA) grants 23MERIT1038415 to A.R. and 24SFRNPCN1284382 and 24SFRNCCN1276092 to A.R. and M.W.

## Author contributions

M.W. conceived the study, supervised the research, and revised the manuscript. A.R. supervised the research. H.Q. performed the analyses and wrote the manuscript. C.W. contributed to the biological interpretation of the findings. B.L. contributed to figure design and layout.

